# Molecular decoupling of lineage identity and morphology in aggressive variant prostate cancer

**DOI:** 10.64898/2026.01.07.26343520

**Authors:** Chennan Li, JuanJuan Yin, Melissa L. Abel, Dana S. Vargas Solivan, Kinjal Bhadresha, Sumeyra Kartal, Samantha Nichols, Kanak Parmar, Joseph Twohig, Tri M. Truong, Cindy H. Chau, Kathleen Kelly, William D. Figg, Anish Thomas, Adam G. Sowalsky

## Abstract

Aggressive variant prostate cancer (AVPC) is a lethal subtype of prostate cancer characterized by its androgen independence, resistance to chemotherapy, and display of neuroendocrine features which can emerge either de novo or via transformation after a prior diagnosis of adenocarcinoma. The poor clinical outcomes in patients with AVPC are associated with its profound molecular heterogeneity. In this study, we analyzed 23 consecutive AVPC cases treated at a dedicated small-cell clinic (2017–2025) using clinicogenomic and transcriptomic profiling. Transformed AVPC exhibited significantly shorter overall survival times than de novo AVPC (11.8 vs 26.0 months, P < 0.001). Integrative genomic analyses identified residual androgen signaling in subsets of cases harboring neuroendocrine lineage programs, highlighting a decoupling of lineage identity and morphology. To facilitate mechanistic and pharmacologic studies, we established NCI-LYM-1, a patient-derived organoid/PDX from an AR-negative, ASCL1+/SYP+ lymph node metastasis, which faithfully recapitulates the donor tumor’s molecular and phenotypic features. Short- and long-read whole-genome sequencing combined with optical genome mapping identified biallelic inactivation of *PTEN*, *TP53*, *RB1* and *BRCA2* as potential drivers, demonstrating clonal concordance with circulating tumor DNA from the original patient donor. Pathway and perturbation analyses suggested that NCI-LYM-1 harbored a strong dependency on apoptotic pathways, which was confirmed by *in vitro* organoid testing with the BCL-2/BCL-xL inhibitor navitoclax (IC_50_: 0.27 µM) and the MCL-1 inhibitor AZD-5991 (IC_50_: 0.060 µM). Overall, NCI-LYM-1 recapitulates the clinical aggressiveness and heterogeneity of AVPC, providing a tractable platform to identify novel precision therapies.

## INTRODUCTION

Although the vast majority of prostate tumors arise from the luminal lineage and remain dependent upon androgen receptor (AR) for survival, a subset of lethal prostate tumors is AR-independent [1, 2]. Broadly termed aggressive variant prostate cancer (AVPC), these tumors harbor neuroendocrine features and often display small cell histologic properties, which widely involve up-regulation of synaptophysin and chromogranin, high proliferation indices, and a low cytoplasm-to-nuclear ratio [3–5]. Notably, these tumors display near or complete indifference to AR-targeted and hormonal therapies. As more potent agents for androgen annihilation are introduced into the clinic for treating patients with metastatic castration-resistant prostate cancer (mCRPC), the incidence frequency of treatment-emergent neuroendocrine prostate cancer has similarly increased [6–8].

While many AVPC cases emerge following transformation of AR-driven prostate cancer adenocarcinoma (ARPC), a rarer variant can emerge de novo in both primary and metastatic settings. These de novo AVPC tumors almost always arise via dedifferentiation or transdifferentiation from previously AR-positive cells and often display anaplastic characteristics [9, 10]. Both de novo and transformed AVPC commonly harbor loss-of-function mutations to *PTEN, TP53*, and/or *RB1* [1, 3, 6, 10]. Irrespective of their origin, however, the treatment options for AVPC are highly limited, with platinum-based chemotherapy and topoisomerase inhibitors being the current standard-of-care [10, 11]. Critically, the clinical outlook for these patients is poor, with median overall survival ranging from 7-16 months depending on the case series [3].

An ongoing challenge towards improving management of AVPC has been the scarcity of tractable model systems that faithfully recapitulate the genomic and phenotypic complexity of human prostate cancer [10, 12]. Although many molecular and mechanistic analyses can be performed directly on biopsy tissue, renewable sources of AVPC models, such as organoid and patient-derived xenografts (PDXs), are required for robust preclinical testing and deep assessment of AVPC tumor biology [13–15]. Here, we hypothesize that integrative molecular profiling of AVPC would uncover lineage-independent therapeutic vulnerabilities and that development of a renewable organoid/PDX model (NCI-LYM-1) would enable mechanistic and preclinical testing. We identify molecular features associated with platinum response in AVPCs from a dedicated small cell cancer clinic. We further demonstrate that a novel organoid/PDX model, isolated from a patient with transformed AVPC with small cell features, faithfully recapitulates the molecular and phenotypic aggressivity of the original tumor while providing essential insight into the distinct pathology of AVPC.

## RESULTS

### Heterogeneous molecular landscape of aggressive variant prostate cancer

The overall goal of this study was to identify molecular and phenotypic properties of aggressive variant prostate cancer (AVPC) that contribute to poor clinical outcomes. We reviewed the clinical outcomes of 23 consecutive patients from 2017 to 2025 with prostate cancer who were referred to our small cell carcinoma clinic for diagnosis of AVPC with neuroendocrine features with or without small cell pathology. When we dichotomized the cohort based on clinical history, overall survival from the time of AVPC diagnosis was significantly shorter in patients who had progressed after prior therapy for prostate adenocarcinoma versus those diagnosed with de novo AVPC (Table 1). Overall median duration of radiographic progression-free survival (rPFS) from the initiation of platinum chemotherapy was 174 days. However, we observed no significant difference in platinum response between de novo (n=11) and transformed (n=12) AVPC (Fig. 1A). Moreover, when we dichotomized the cohort based on the presence (n=11) or absence (n=12) of small cell pathologic features identified on biopsy, no significant rPFS difference was observed either (Fig. 1B). All of these patients experienced rPFS within 13 months.

**Figure 1.**
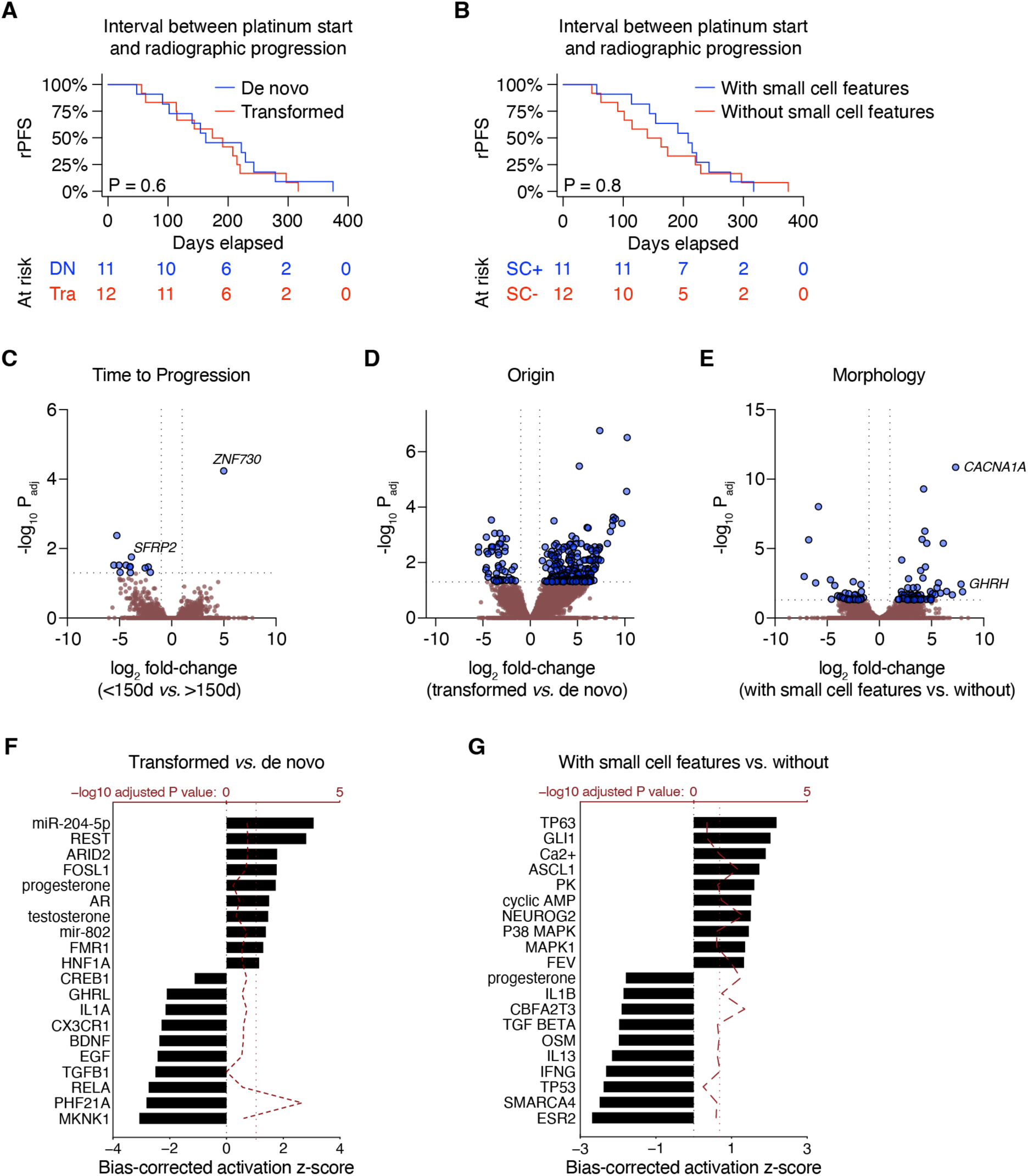
Molecular differences amongst aggressive variant prostate cancers. **(A–B)** Stratification of the AVPC cohort based on de novo vs. transformed AVPC **(A)** and AVPC with vs. without small cell features **(B)**. P values are determined using log-rank test. **(C–E)** Volcano plots depicting differentially-expressed genes (DEGs) comparing radiographic progression before or after the mean rPFS of 150 days **(C)**, transformed *vs.* de novo AVPC **(D)**, and with *vs.* without small cell features **(E)**. Horizontal boundary depicts the *P* = 0.05 (adjusted) cutoff. **(F–G)** All statistically significant DEGs from **D–E**, respectively (*P*_adj_ < 0.05) were processed with the upstream regulator module of Ingenuity Pathway Analysis. The ten most activated and inactivated pathways (with adjusted *P* values less than 0.05) are shown. The bias-corrected *z* score is shown on the bottom *x*-axis and the adjusted *P* value is shown on the top *x*-axis (-log_10_ transformed).

**Table 1.** Patient characteristics of the NCI AVPC cohort. ADPC: prostate adenocarcinoma; AVPC: aggressive-variant prostate cancer. ^a^ Welch’s *t* test. ^b^ Fisher’s exact test. ^c^ Log-rank test.

| Demographics (N=23) |  | De novo (n=11) | Transformed (n=12) | P value |
| --- | --- | --- | --- | --- |
|  |  | Median (IQR) | Median (IQR) |  |
| Age at ADPC diagnosis (y) |  | N/A | 59 (54–68) | N/A |
| Age at AVPC diagnosis (y) |  | 69 (55–72) | 64 (60–76) | 0.9<sup>a</sup> |
| Race |  | % (N) | (N) |  |
| Black |  | 0 | 33 (4) |  |
| Asian |  | 0 | 0 |  |
| White |  | 82 (9) | 67 (8) |  |
| Unknown |  | 18 (2) | 0 |  |
| **Pathology** |  |  |  |  |
| Initial Gleason score (grade group) |  | % (N) | % (N) | 1<sup>b</sup> |
| 3+3 (1) |  | 9 (1) | 17 (2) |  |
| 3+4 (2) |  | 0 | 8 (1) |  |
| 4+3 (3) |  | 0 | 17 (2) |  |
| 4+4; 3+5; 5+3 (4) |  | 0 | 0 |  |
| 4+5; 5+4; 5+5 (5) |  | 27 (3) | 50 (6) |  |
| Small cell features present |  | 36 (4) | 58 (7) | 0.4<sup>b</sup> |
| **Clinical** |  |  |  |  |
|  |  | Median (IQR) | Median (IQR) |  |
| PSA on ADPC diagnosis (ng/mL) |  | N/A | 6.6 (5.7–14.6) | N/A |
| PSA on AVPC diagnosis (ng/mL) |  | 2.4 (0.9–5.3) | 0.1 (0–2.5) | 0.3<sup>a</sup> |
| Overall survival since ADPC diagnosis (m) |  | N/A | 41.2 (32.8–102.0) | N/A |
| Transformation interval (m) |  | N/A | 20 (8–25) | N/A |
| Overall survival since AVPC diagnosis (m) |  | 26.0 (16.5–30.6) | 11.8 (10.1–19.1) | < 0.001<sup>c</sup> |
|  |  | % (N) | % (N) |  |
| Metastatic disease at first diagnosis |  | 82 (9) | 33 (4) | 0.036<sup>b</sup> |
| Bone-dominant metastases |  | 27 (3) | 42 (5) | 0.7<sup>b</sup> |
| Prior androgen blockade |  | 36 (4) | 92 (11) | < 0.01<sup>b</sup> |
| Prior androgen receptor pathway inhibitor |  | 18 (2) | 75 (9) | 0.012<sup>b</sup> |
| Prior platinum |  | 100 (11) | 75 (9) | 0.2<sup>b</sup> |
| Prior etoposide |  | 100 (11) | 67 (8) | 0.093<sup>b</sup> |
| Prior atezolizumab |  | 27 (3) | 33 (4) | 1<sup>b</sup> |
| Prior taxane |  | 9 (1) | 33 (4) | 0.3<sup>b</sup> |
| Prior lurbinectedin |  | 9 (1) | 8 (1) | 1<sup>b</sup> |
| Partial response to platinum |  | 73 (8) | 58 (7) | 0.7<sup>b</sup> |

Of these 23 patients, 9 had evaluable biopsies suitable for RNA extraction and subsequent whole-transcriptome sequencing. Strikingly, there were very few differentially-expressed genes (DEGs) distinguishing those with earlier radiographic progression (less than the mean, 150 days) versus those who progressed later (Fig. 1C). Far more DEGs were identified when comparing de novo vs. transformed AVPC (Fig. 1D) and with vs. without small cell features (Fig. 1E). Upstream regulator analysis of these DEGs revealed residual AR activity enriched in transformed AVPCs (Fig. 1F) and greater representation of neuronal lineage drivers NEUROG2 and ASCL1 (Fig. 1G).

To further assess genomic and genetic differences between AVPC tumors and other lethal metastatic castration resistant prostate cancers (mCRPCs), we analyzed exome sequencing data from 12 evaluable AVPC biopsies from which DNA was obtained and integrated these data with whole-genome/exome somatic copy number estimates from 20 other tissues/PDX/PDO samples representing various mCRPC subtypes (16 LuCaP-series PDX or PDX-O samples, two PDO samples from NCI, plus two biopsies from the Prostate Cancer Foundation Stand Up to Cancer West Coast Dream Team (WCDT) cohort). Arm level gains, which are amongst the frequently altered genomic events in early prostate cancer, were equivalently distributed between AVPC and AR-positive (ARPC) tumors (Fig. 2A). At the gene level, biallelic hits to *TP53* and *RB1* were overrepresented in AVPC (55% and 45%, respectively), and no *AR* point mutations were observed (Fig. 2B).

**Figure 2.**
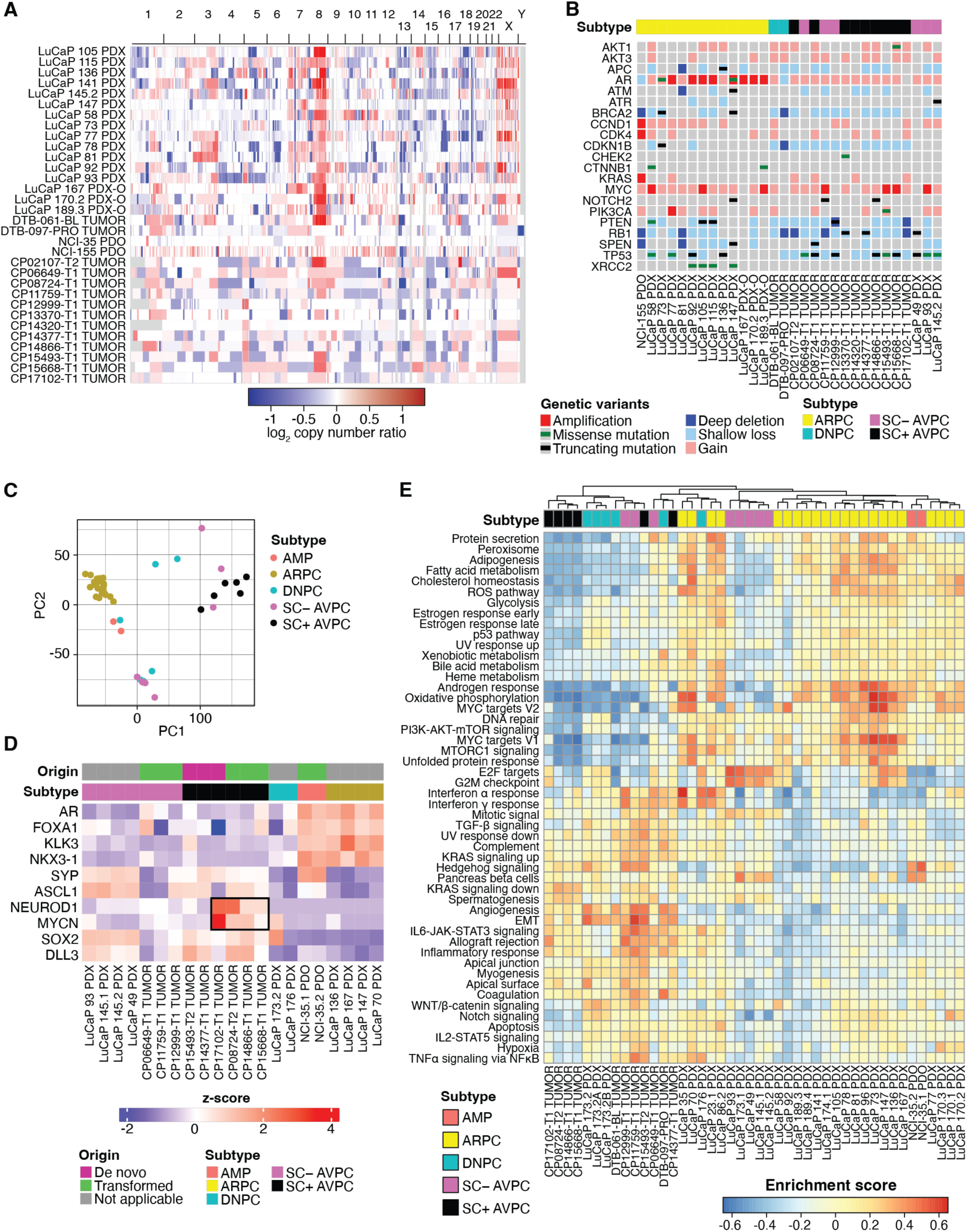
Molecular landscape of aggressive variant prostate cancers. **(A)** Whole-genome somatic copy-number estimates derived from whole-exome or whole-genome sequencing of prostate tumor patient-derived xenograft (PDX), PDX-to-organoid (PDX-O), and biopsy samples. **(B)** Oncoprint depicting copy number and mutational status of selected prostate cancer genes altered in at least one AVPC case. All mutations shown were curated for known oncogenic status. **(C)** Principal component analysis of 45 mCRPC tumor transcriptomes, colored by subtype. **(D)** Heatmap depicting ARPC and AVPC lineage-defining gene expression by *z*-score. **(E)** Heatmap depicting unsupervised nonparametric gene set variation analysis for 45 mCRPC tumor transcriptomes projected against the mSigDB Hallmarks gene sets.

To evaluate the transcriptomic characteristics of AVPC tumors, bulk RNA-seq data from 45 mCRPC sources (12 AVPC biopsies from our study, 31 LuCaP-series PDX/PDX-O samples, two PDO from NCI, and two biopsies from WCDT). We used principal component analysis (PCA) to evaluate global differences between AVPC and other mCRPC tumors. Despite making further distinctions as to small cell or neuroendocrine subtypes, most AVPC shared transcriptional similarities with each other (Fig. 2C). Closer examination of lineage-defining genes for ARPC (*AR*, *FOXA1*, *KLK3*, and *NKX3-1*) and AVPC (*SYP*, *ASCL*, *NEUROD1*, *MYCN*, *SOX2* and *DLL3*) demonstrated that a subset of AVPC (specifically, transformed with small cell features) expressed high levels of *NEUROD1* and *MYCN* exclusive of neuroendocrine feature-positive prostate cancer (NEPC) and most de novo AVPCs (Fig. 2D). To identify cellular pathways that were enriched amongst AVPC subtypes, an unsupervised nonparametric gene set variation analysis (GSVA) was conducted. Consistent with the PCA result, we observed clustering among SC+ AVPC, SC− AVPC, and double-negative prostate cancer (DNPC) which consistently displayed lower androgen response, metabolism, and DNA repair than ARPC and amphicrine prostate cancer (AMP). By contrast, most AVPCs were characterized by greater enrichment for cell cycle checkpoint, inflammatory signaling and epithelial-to-mesenchymal transition (EMT) (Fig. 2E). Collectively, these data point to profound molecular heterogeneity amongst mCRPC with distinctive aggressive phenotypes associated with AVPC tumors.

### Generation of a novel AVPC model transformed from ARPC with small cell features

As part of ongoing efforts to assess the biology underlying poor clinical outcomes for patients with AVPC, we generated an organoid-to-PDX model from patient CP-08724, for whom biopsy samples had been previously acquired of his transformed AVPC harboring small cell features (see Fig. 2D). The tumor biopsy specimen was acquired from a Black man (CP-08724), previously diagnosed with low-risk prostate adenocarcinoma (Gleason score 3+3) in his 50s, and had been treated with leuprolide and brachytherapy which had successfully reduced prostate-specific antigen (PSA) (Fig. 3A). He experienced biochemical recurrence approximately five years after initial diagnosis and was found to have small equivocal bone lesions on PSMA-PET that were not clearly seen on a conventional bone scan. He was then treated with three cycles of Radium-223 as part of a clinical trial, during which he experienced disease progression with increasing metastasis seen on conventional bone scan (Fig. 3B). PSA levels dropped in response to subsequent androgen deprivation therapy (goserelin/degarelix) plus docetaxel chemotherapy. Despite PSA levels remaining low, however, he experienced rapid progression with new visceral metastasis on CT scans (Fig. 3C). Biopsy-confirmed metastatic lesions in the bladder, rectum, and bone were observed with a pathologic diagnosis of prostate cancer with small cell features, which was supported by negative PSA, positive synaptophysin (SYP) and positive chromogranin (CHGA) on biopsy tissue immunohistochemistry (IHC) (Fig. 3D). He then received a standard-of-care neuroendocrine prostate cancer treatment regimen with chemo- and immuno-therapies (carboplatin, etoposide, and atezolizumab) extrapolated from small cell lung regimens. A partial response was observed after four cycles of treatment, with radiographic progression subsequently observed in the liver. This patient was then treated in a dedicated small cell cancer clinic with an ATR inhibitor (berzosertib) in combination with a Trop-2 antibody and topoisomerase inhibitor conjugate (sacituzumab govitecan) and subsequently with berzosertib in combination with a standard second-line small cell chemotherapy (topotecan) as part of a clinical trial (see Fig. 3A). However, only brief responses were observed from either treatment.

**Figure 3.**
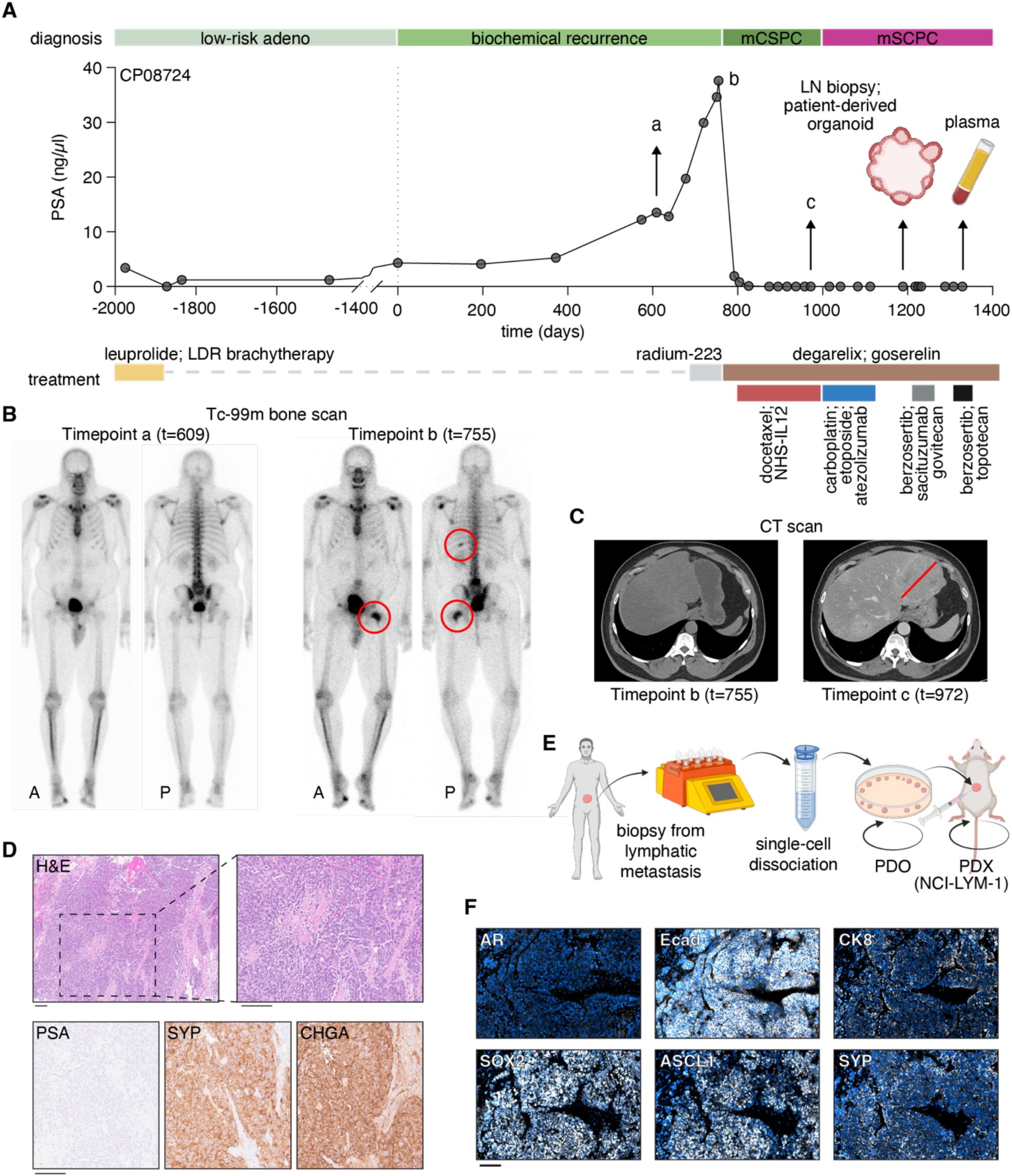
Clinical history and derivation of the NCI-LYM-1 patient-derived AVPC model. **(A)** Serum PSA trajectory for patient CP-08724, with each point indicating a blood draw. Clinicopathologic disease state is given at the top with time on the *x*-axis. Single or overlapping treatments are indicated by horizontal bars beneath the *x*-axis. a, b, and c denote timepoints of diagnostic imaging. **(B)** Bone scans at timepoints a and b, with red circles identifying lesions indicative of potential metastatic growth. **(C)** Computed tomography (CT) scans at timepoints b and c identifying development of visceral metastases. **(D)** H&E and immunohistochemistry (IHC) staining of serial diagnostic sections taken at timepoint c that indicate the development of neuroendocrine features. Bar: 100 µm. PSA: prostate-specific antigen; SYP: synaptophysin; CHGA: chromogranin A. **(E)** Schematic depicting the establishment of the NCI-LYM-1 patient-derived organoid model (PDO) that was subsequently grown subcutaneously as a patient-derived xenograft (PDX). **(F)** Immunofluorescent staining of subcutaneous tumors stained with antibodies against androgen receptor (AR), E-cadherin (Ecad), cytokeratin 8 (CK8), SRY-box 2 (SOX2), Achaete-Scute homolog 1 (ASCL1), and SYP. Bar: 100 µm.

After platinum chemotherapy but prior to starting berzosertib combination therapy, we received a lymph node metastasis biopsy from the patient for generating a renewable research model. This model, which we have termed NCI-LYM-1, was developed following our established practices for single-cell dissociation of tissue [14–16], growth as a patient-derived organoid (PDO) and subsequent expansion as a patient-derived xenograft (PDX) (Fig. 3E). Subcutaneous tumors displayed phenotypes consistent with biopsy IHC, including negative staining for AR and positive staining for ASCL1, SOX2, and SYP (Fig. 3F). Notably, these tumors also expressed E-cadherin and retained expression of cytokeratin 8 (CK8) (Fig. 3F), confirming their epithelial origin as prostate cancer tissue.

### Identifying molecular and phenotypic drivers of AVPC via integrative structural genomics

Given the histological similarities between the NCI-LYM-1 model and its originating tumor tissue, we next sought to use this renewable resource for establishing a genomic basis for disease progression and treatment resistance, starting with three different forms of whole-genome sequencing applied to genomic DNA (gDNA) isolated from NCI-LYM-1 PDX cells and matched buffy coat gDNA. Germline WGS-based ancestry was estimated at 73.5% African and 26.5% European (see methods). As depicted in Figure 4A, we integrated conventional short-read (Illumina) sequencing at 152× coverage, long-read (Oxford-Nanopore Technologies, ONT) sequencing at 112× coverage, and optical genome mapping (OGM, Bionano) at 600–700× coverage. In addition to confirming the presence of the *TP53* frameshift mutation we had previously observed in the donor biopsy tissue (Supplementary Fig. 1A), we successfully phased ONT reads into two distinct haplotypes to identify monoallelic losses to *TP53* and *PTEN,* and biallelic inactivation of *RB1*, being the major genomic drivers (Fig. 4B). Using OGM, we resolved that hits to *RB1* consisted of a genomic deletion coupled with a translocation to chromosome 19 that interrupted the reading frame (Fig. 4C).

**Figure 4.**
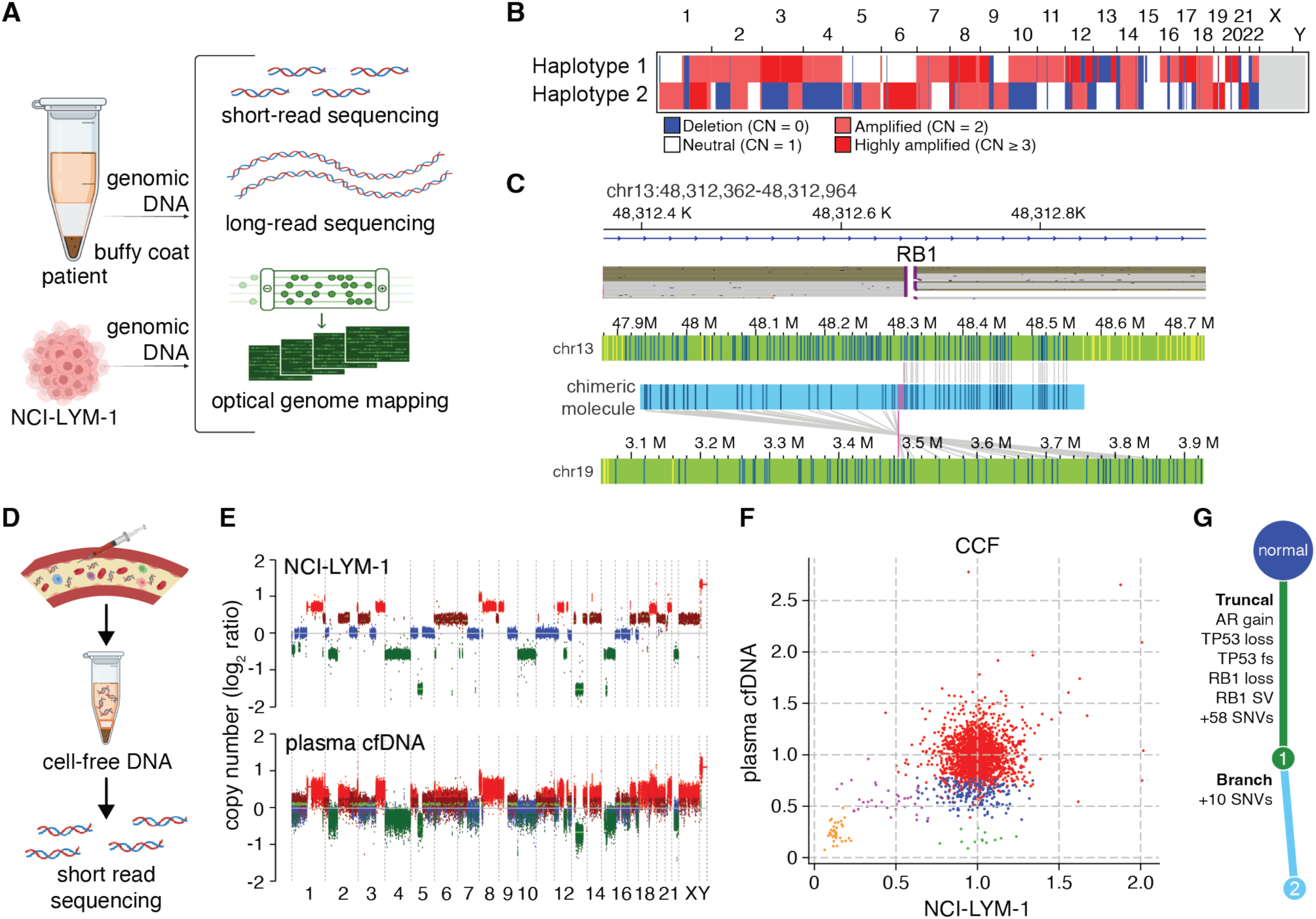
Comparison of molecular features between PDX and original tumor. **(A)** Schematic showing an integrative sequencing approach for whole-genome profiling described in this and subsequent figures. **(B)** Genome-wide copy number profile of each haplotype in NCI-LYM-1 PDX as resolved by long-read sequencing. **(C)** Top: long-read sequence reads of high-molecular weight DNA from NCI-LYM-1 PDX tissues, mapping breakends within *RB1* as visualized in the Integrative Genome Viewer. Bottom: optical genome mapping of ultra-high molecular weight DNA from NCI-LYM-1 PDX tissue, visualizing translocations between *RB1* on chromosome 13 and chromosome 19. **(D)** Schematic showing the whole-genome short-read sequencing approach described in this and subsequent figures. **(E)** Genome-wide somatic copy number variation profiles of NCI-LYM-1 genomic DNA (top) and patient CP-08724 circulating cell-free tumor DNA (bottom). **(F)** Common and private somatic mutations are represented as cancer cell fractions (CCF) from patient CP-08724 circulating cell-free tumor DNA (*y*-axis) and NCI-LYM-1 (*x*-axis). Each group of colored dots indicates distinct clones. The truncal clone has a CCF of ∼1, while subclones have lower CCFs. **(G)** Reconstruction of phylogenetic architecture using all available genomic structural variant, copy number, and mutational data places multiple driver events as clonal/truncal perturbations (green) and passenger variants as a subclone/branch (light blue).

PDX and PDO models like NCI-LYM-1 were established from a subset of tumor cells from a biopsy sample, which itself is a subset of systemic tumor burden. To compare the clonal composition and cellular heterogeneity between NCI-LYM-1 and the lethal tumor from patient CP-08724, we performed 155× short-read sequencing on circulating cell-free DNA (cfDNA) (Fig. 4D) isolated from plasma drawn during a subsequent clinical visit (see Fig. 3A) as a genomic readout of systemic tumor burden [17]. Compared to buffy-coat short-read sequencing (at 123× coverage), genome-wide patterns of somatic copy number variation (SCNV) were nearly identical between NCI-LYM-1 and plasma cfDNA (Fig. 4E and Supplementary Fig. 1B). However, computational modeling of cancer cell fractions [18] of individual point mutations indicated differences in clonal architecture. Whereas both NCI-LYM-1 and plasma cfDNA shared a single apparent major clone with two minor subclones, plasma cfDNA harbored at least two additional minor clones that had reached ∼100% clonality in the PDX (Fig. 4F). Integrating these findings together, we resolved a highly similar clonal composition to the donor tumor, with all previously recognized driver mutations being present in every cell of NCI-LYM-1 (Fig. 4G).

Having used a PCR-free approach (see methods) to generate our ONT libraries, we called methylated DNA bases from PDX and buffy coat long-read sequencing libraries for inferring gene silencing due to promoter and enhancer methylation (Fig. 5A). Phasing in our existing SCNV data, we found that the remaining allele of *PTEN* was hypermethylated, which combined with genomic findings indicated total biallelic inactivation of *PTEN* (Fig. 5B). We noted additionally a biallelic inactivation of *BRCA2*, supported both by loss-of-heterozygosity (LOH, consistent with prior patient biopsy sequencing, see Fig. 2B) and hypermethylation of the remaining allele (Fig. 5C), a feature not evident from prior sequencing. Distinctly calling 5′-hydroxymethylcytosine (5hmC) and 5′-methylcytosine (5mC) modifications, we further inferred genome-wide patterns of silencing to identify potential active transcription factors based on known transcription factor binding sites (TFBSs) of active genes (Fig. 5D). Amongst the TFBSs with the lowest levels of 5mC or highest levels of 5hmC (compared to buffy coat) were those associated with ASCL1 activity, a well-characterized driver of AVPC (Fig. 5E), OSR2 which is known to regulate anti-tumor lymphocyte activities [19], FEZF1 which is involved in nervous system development [20], and GRHL2 which regulates EMT [21]. *ZBTB16* (PLZF), a transcription factor (TF) lost in CRPC [22], was amongst the top TFs putatively depleted for 5hmC and enriched for 5mC at their binding sites in NCI-LYM-1.

**Figure 5.**
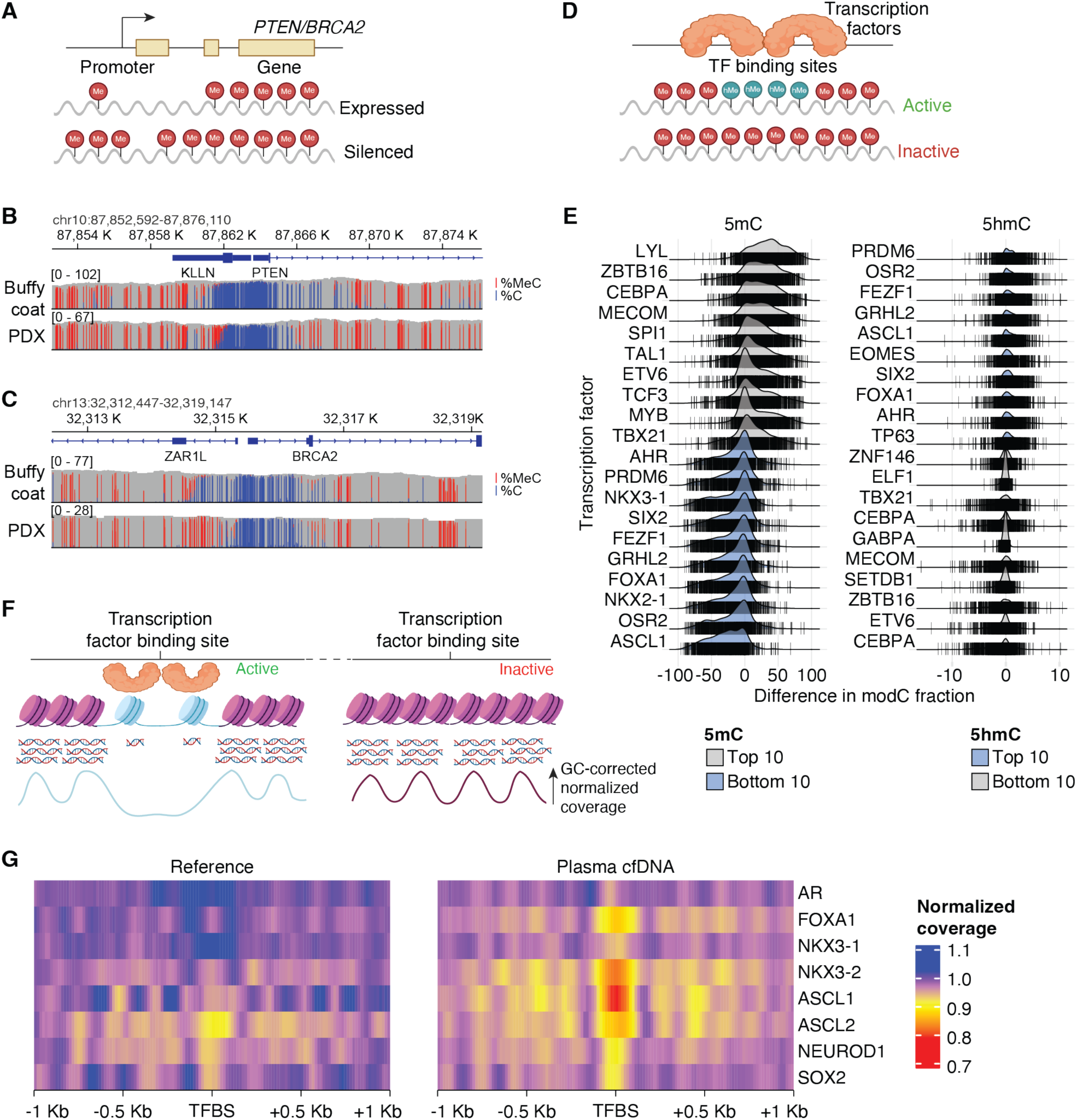
Inference of phenotypic and transcriptomic alterations using long-read NCI-LYM-1 tissue and short-read patient CP-08724 cell-free DNA sequencing. **(A)** Schematic showing the inference of gene silencing using DNA methylation from long-read sequencing. (**B–C)** IGV tracks showing the differences in proportions of reads methylated (in red) for the promoter and the first introns of *PTEN* (B) and *BRCA2* (C) between the NCI-LYM-1 PDX (bottom) and matched buffy coat from the donor, CP-08724 (top). **(D)** Schematic depicting the activation or repression of gene expression at known transcription factor binding sites (TFBSs) based on long-read sequencing 5′-methylcytosine (5mC) and 5′-hydroxymethylcytosine (5hmC) base calls, respectively. **(E)** Ridgeline plots showing the differentially methylated TFBSs determined by 5mC and 5hmC base calls from long-read tissue sequencing compared to the matched buffy coat. Differences at each TFBS are shown as vertical lines that cross the horizontal line. The top 10 and bottom 10 differentially enriched TFs inferred are shown. TFs are ordered based on the median difference of 5mC or 5hmC. **(F)** Schematic showing the molecular basis for inferring transcriptional activity based on fragmentation patterns of cell-free DNA deprotecting open chromatin (top) and GC-corrected normalized cfDNA coverage (bottom). **(G)** GC-corrected normalized cfDNA coverage across each TFBS ± 1Kb is shown. Reduced depths indicative of open chromatin are shown in yellow/red. A ctDNA-negative plasma sample was used as the reference.

Finally, we examined patterns of cfDNA fragmentation (Fig. 5F), which have been shown to accurately predict nucleosome occupancy genome-wide and therefore chromatin accessibility and activity of transcription factors for the purpose of predicting gene expression [23]. Consistent with inferred TF activity, ASCL1 had the greatest reduced GC-corrected normalized coverage at its TFBSs in plasma cfDNA, implying active transcription (Fig. 5G). NKX2-1 (another NEPC marker [24]) also showed strong activity, while the adenocarcinoma lineage transcription factors AR and NKX 3.1 indicated far less inferred transcriptional activity (Fig. 5G). Collectively, these data demonstrate faithful recapitulation of the genomic complexity of AVPC in the NCI-LYM-1 PDX model.

### Profiling of pharmacological and metastatic phenotypes of the NCI-LYM-1 model

Notwithstanding the lack of apparent platinum response differences between transformed and de novo AVPCs and AVPCs with and without small cell features (see Fig. 1A–B) we further extrapolated potential drug sensitivities in transformed and small cell-positive AVPCs by applying the differentially expressed gene lists (from Fig. 1D–E) to the Connectivity Map (Fig. 6A–B). The topmost hit for predicted sensitivity in transformed AVPCs (out of 35,811 total perturbations) was the BH3 mimetic navitoclax, which we validated to have moderate activity in NCI-LYM-1 organoid culture at an IC_50_ of 0.27 µM (Fig. 6C). Another BH3 mimetic targeting MCL-1, AZD-5991, showed even stronger activity with an IC_50_ of 0.060 µM (Fig. 6D). The drug predicted to have least activity amongst this group was foretinib, a pan-receptor tyrosine kinase inhibitor, and indeed ipatasertib (AKT inhibitor) showed weak activity against NCI-LYM-1 at 1.9 µM (Fig. 6E).

**Figure 6.**
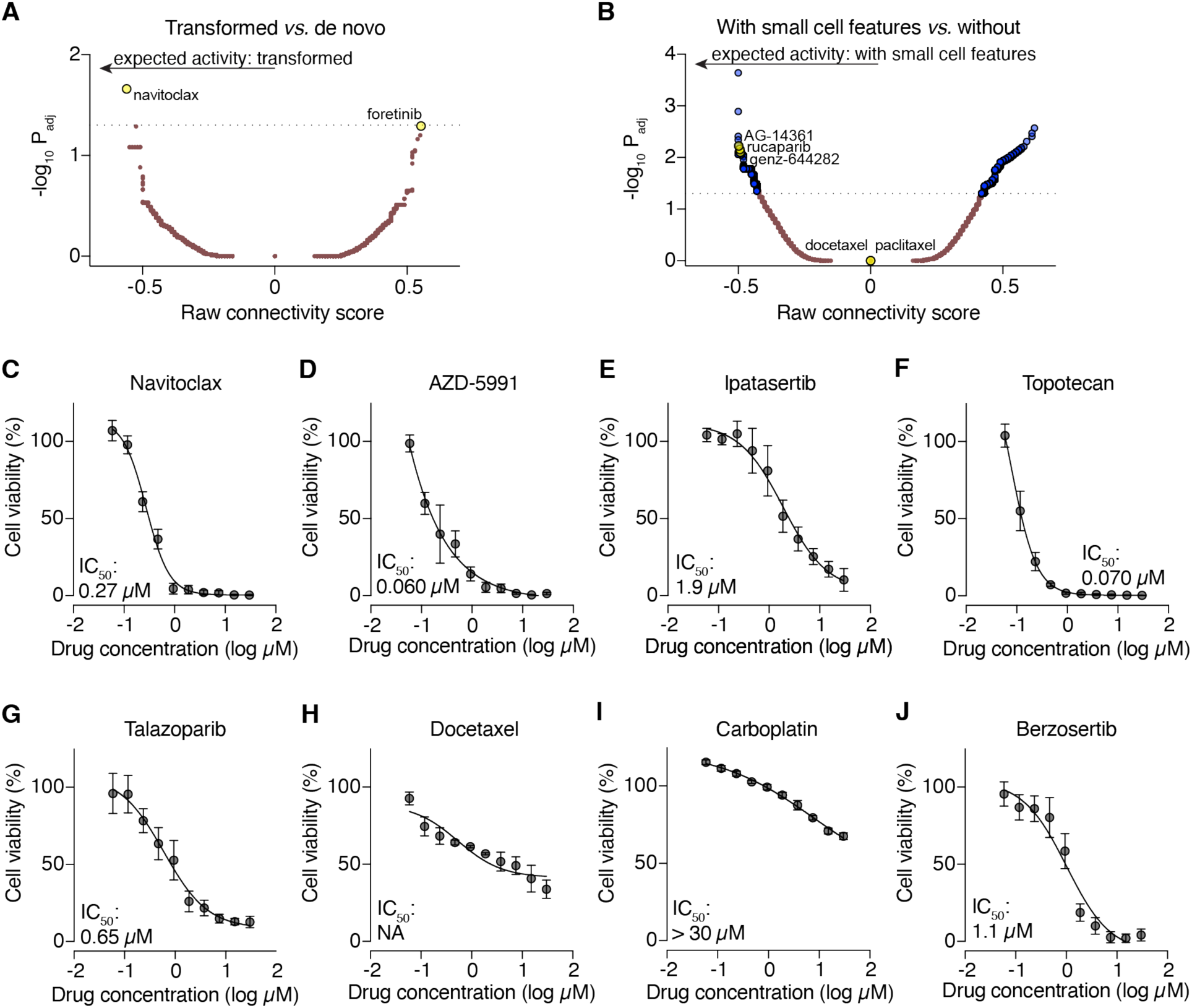
Evaluation of AVPC targeting agents. **(A–B)** The top differentially-expressed genes comparing transformed *vs.* de novo AVPC (A) and AVPC with *vs.* without small cell features (B) were processed with Connectivity Map. The raw connectivity scores and adjusted P values are shown as a volcano plot. **(C–J)** Dose-response curves (DRC) of NCI-LYM-1 organoid culture viability assays following treatment with small-molecule targeted agents or chemotherapies as informed by Connectivity Map scores or clinical behavior of the patient donor, CP-08724. Each viability assay was performed three times independently.

The comparison of AVPCs with and without small cell features yielded many more potential hits, with the topoisomerase inhibitor genz-644282 being amongst the top 0.1% of Connectivity Map hits for SC+ AVPCs. Given that the NCI-LYM-1 patient donor had no prior exposure to topoisomerase inhibitors, the TOP1 inhibitor topotecan displayed potency at 0.070 µM (Fig. 6F). Consistent with biallelic inactivation of *BRCA2* (see Figs. 2B and 5B), predicted sensitivity with e PARP inhibitors (such as AG-14361 and rucaparib) was also confirmed *in vitro* using talazoparib (0.65 µM, Fig. 6G). As the patient donor for NCI-LYM-1 had previously progressed on docetaxel and carboplatin (see Fig. 3), neither had strong antitumor activity *in vitro* (Fig. 6H–I), which also aligned with computationally predicted behavior for both docetaxel and paclitaxel. Finally, owing to the fact that the patient exhibited brief responses to berzosertib (ATR inhibitor, see Fig. 3A), we assessed its IC_50_ *in vitro*. NCI-LYM-1 displayed only weak sensitivity to berzosertib (1.1 µM, Fig. 6J). Taken together, these data demonstrate that NCI-LYM-1 displayed a complex phenotype of drug responses consistent with the donor patient’s clinical history.

The aggressive properties of AVPC are due in part to its tendency to metastasize [3]. We therefore proceeded to assess this model’s metastatic potential using an intracardiac injection approach (Fig. 7A). To monitor disease progression *in vivo*, prior to inoculation, NCI-LYM-1 cells were transduced with lentiviral particles to express luciferase. Bioluminescent intensity (BLI) was detected 5 weeks after inoculation (Fig 7B). Metastases established in 92% (11/12) mice, with tumor burden (as measured by BLI) doubling every 3–4 days (Fig. 7C) and reaching ethical endpoints 7–15 weeks after inoculation (Fig. 7D).

**Figure 7.**
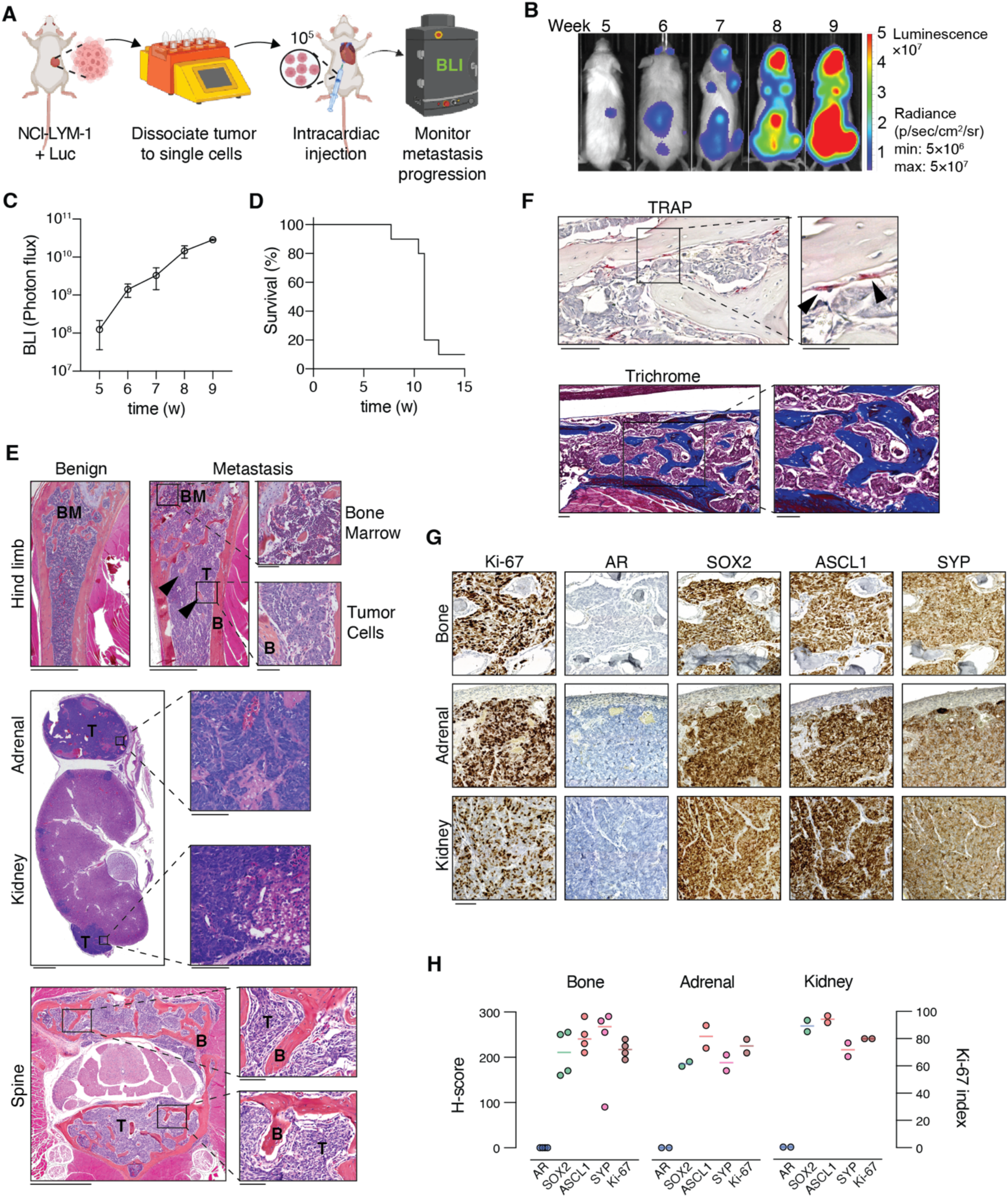
*In vivo* metastatic phenotypes of NCI-LYM-1 recapitulate clinical behavior. **(A)** Schematic showing the workflow for intracardiac injection. **(B)** Representative images (n=5) of weekly bioluminescence (BLI) measurements following inoculation of NCI-LYM-1. **(C)** Quantified whole-body BLI over time. **(D)** Survival rate of NCI-LYM-1 bearing mice upon reaching ethical endpoints. **(E)** H&E stains of representative metastases in the long bone, kidney, adrenal and spine. Arrowheads: new bone. T: tumor; B: bone; BM: bone marrow. Bar: 1 mm; inset bar: 100 µm. **(F)** Top: tartrate-resistant acid phosphatase (TRAP) staining of metastases to long bone. Arrowheads: osteoclasts. Bar: 100 µm; inset bar: 50 µm. Bottom: Masson’s trichrome staining of metastases to long bone. Bar: 100 µm. Arrowheads: new bone. **(G)** Representative immunohistochemistry with antibodies against Ki-67, AR, SOX2, ASCL1 and SYP in serial sections of metastases to the bone (top), adrenal (middle) and kidney (bottom). Bar: 100 µm. **(H)** Whole slide series of tissues shown in (G) were subjected to semi-quantitative H-scoring (left *y*-axis) or Ki-67 index (right *y*-axis). Bar at median.

As shown in Figure 7E, tumor-bearing mice developed metastases in both bone and soft tissues, with robust tumor cell growth in the long bones, kidney, adrenal gland and spine. In the bone, both osteolytic and osteoblastic activities were observed as detected by TRAP and trichrome staining, respectively (Fig. 7F). Immunostaining of metastatic IHC tissues showed patterns of Ki-67, AR and SYP (Fig. 7G) staining comparable to both diagnostic tissues and subcutaneous tumors (see Fig. 3), confirming the stability of the neuroendocrine phenotype. Semi-quantitative H-scoring did not reveal appreciable differences between the staining levels amongst bone, adrenal and kidney metastases (Fig. 7H), with all three sites harboring highly proliferative (70–80%) tumors. Similar to subcutaneous tumors, these metastases also had high levels of SOX2, ASCL1, and SYP staining that did not vary between metastatic sites (Fig. 7G–H). These data establish that the NCI-LYM-1 model recapitulates robust aggressive properties of lethal AVPC.

## DISCUSSION

Rapid disease progression in patients with AVPC has impeded the development and testing of new therapies, such that median overall survival (OS) from the time of AVPC diagnosis has not improved substantially over the last thirty years [1, 5, 25]. In our case series, in which patients progressing on platinum were subsequently enrolled into trials for ATR and topoisomerase inhibitors, median OS for transformed AVPC was less than one year (see Table 1). Gene expression derived from these biopsies inferred sensitivity to BH3 mimetics, and our novel PDO/PDX model, NCI-LYM-1, faithfully recapitulated this drug response (see Fig. 6). Importantly, NCI-LYM-1 also demonstrated similar growth and metastatic properties as the original tumor, highlighting its reliability as a paradigm for improving precision medicine in this critical patient population.

NCI-LYM-1 harbored biallelic loss-of-function events for *PTEN*, *TP53*, and *RB1*, the most frequently silenced tumor suppressors in AVPC [26, 27], with the first allele of all three genes being lost via loss-of-heterozygosity, the most common pattern of sequential haploinsufficiency in prostate cancer [28, 29]. Given the relative importance of *PTEN* to castration-resistant phenotypes and *RB1* to AVPC development [26, 30], the second hits to these genes would have been missed if we had relied on exome sequencing entirely (see Figs. 2B and 5B and Supplementary Fig. 1A); whole-genome and epigenetic approaches were necessary to resolve those additional alterations to the remaining alleles, respectively. Similarly, sensitivity to PARP inhibitors could have been inferred by both genomic and epigenetic modifications to *BRCA2*. Although we describe only a handful of genomic assays, the renewable cellular resource afforded by models such as NCI-LYM-1 increases opportunities for further functional and mechanistic analyses using both *in vitro* and *in vivo* approaches.

Remarkably, our analyses revealed that NCI-LYM-1, which was isolated from a lymph node biopsy, was highly similar in clonal composition to the systemic tumor at the time of sample acquisition. Plasma-derived circulating tumor DNA isolated six months later was highly concordant for somatic copy number alterations and point mutations (see Supplementary Fig. 1A–B). NCI-LYM-1 harbored all major and minor clones detectable in ctDNA (see Fig. 4F), making NCI-LYM-1 representative of the patient donor’s intratumoral heterogeneity. This clonal complexity improves the suitability of the model for preclinical testing, better mimicking the heterogeneous responses of actual tumors to novel therapies.

As a new model we are making available to the prostate cancer community, NCI-LYM-1 is notable as an organoid/PDX derived from a man of African descent. In the United States, Black men have both highest incidence rate of prostate cancer and the greatest risk of dying from prostate cancer [31, 32]. The molecular landscapes of prostate tumors in men of African descent differ sharply from men of European descent, indicating that these apparent ethnic differences are further grounded by genetic factors that transcend racial and other socio-economic considerations [33, 34]. Given that most cell line and PDX models for advanced prostate cancer were derived from tumors from White men [35], NCI-LYM-1 may contribute to expanding the repertoire of African ancestry prostate cancer cell lines that are available for developing more effective treatments.

An important limitation of this study is that future experimentation will be needed to understand the biologic implications of genomic and phenotypic observations. Similar to the donor tissue, NCI-LYM-1 showed similar drug resistance profiles and highly aggressive metastatic properties *in vivo* (see Figs. 6 and 7). The model also displayed elevated expression of proliferation-associated genesets (E2F targets, G2M checkpoints, Supplementary Fig. 1C), findings consistent with other AVPC PDX models and with the uniformly high Ki-67 indices in NCI-LYM-1 PDX metastatic lesions (see Figs. 2E and 7G, respectively). Moreover, NCI-LYM-1 is ideally suited to tease apart the association of small cell morphology and molecular features, such as expression of *NEUROD1/MYCN* or *ASCL1*, and to test whether epigenomic perturbation using EZH2, BET, or HDAC inhibitors can influence small cell morphology *in vivo*. Thus, the NCI-LYM-1 model offers a robust platform for future mechanistic investigation to assess gene-level dependencies for maintaining these aggressive and lethal properties in AVPC.

Similar to efforts performed using other PDX and PDO models [36], NCI-LYM-1 demonstrated utility for drug screening of novel therapies in AVPC in which models harboring distinct molecular phenotypes may be suited to guide future clinical decision-making for drug sensitivity testing. For example, NCI-LYM-1 recapitulated the patient’s own responses as measured by resistance of the PDO to docetaxel and carboplatin. By contrast, the strong activity by AZD-5991 (MCL-1 inhibitor) represents a novel approach for targeting transformed AVPC tumors including NCI-LYM-1. Indeed, we recently reported that *MCL1* genomic gains in CRPC are associated both with poorer clinical prognosis and greater sensitivity to MCL-1 inhibitor monotherapy [37], and NCI-LYM-1/CP08724 was amongst 5 AVPC tumors from the NCI cohort with 1q21 gains that encompassed *MCL1*. In addition, our organoid drug screening also revealed topotecan as highly effective in this model. In line with the recent report that metronomic topotecan alone or in combination with docetaxel could suppress EMT and stem-like CD44^+^/CD133^+^ populations [38], our findings suggest that topotecan plus docetaxel may inhibit AVPC progression by targeting stemness, EMT programs and key oncogenic pathways. Thus, NCI-LYM-1 could serve as a precision pharmacology platform to identify and test novel agents to treat AVPC.

The lack of durable therapies for AVPC represents an unmet clinical need. This work establishes NCI-LYM-1 as a renewable, genomically annotated resource that recapitulates response patterns for a distinct molecular subtype of AVPC, and our data collectively suggest that NCI-LYM-1 amongst other platforms will be a valuable tool for identifying new therapeutic targets. We anticipate that its continued use will inform mechanism-guided trials and improve outcomes for patients with AVPC.

## PATIENTS AND METHODS

### Study Approval

This trial was approved by the Institutional Review Board of the Center for Cancer Research, NCI (Bethesda, MD; ClinicalTrials.gov Identifier: NCT04826341, first registered on March 31, 2021). This study was conducted in accordance with ethical principles that have their origin in the Declaration of Helsinki and are consistent with the International Council on Harmonization guidelines on Good Clinical Practice, all applicable laws and regulator elements, and all conditions required by a regulatory authority and/or institutional review board. Written informed consent was obtained from all patients prior to performing study-related procedures in accordance with federal and institutional guidelines.

### Biosample acquisition

CT-guided biopsies were acquired by an interventional radiologist to sites of tumor previously identified on MRI, CT, FDG-PET or PSMA-PET prior to the initiation of therapy or upon radiographic progression. Biopsies for diagnostic use were fixed in formalin and embedded into paraffin following standardized practices. All diagnoses of neuroendocrine carcinoma and/or small cell features were confirmed by board-certified NIH pathologists.

To generate the NCI-LYM-1 organoid, a lymph node metastasis biopsy sample was processed immediately after acquisition with the protocol described previously [14]. 2 × 10^5^ cells in Hans Clevers media [14] (HCM) containing 80% of Matrigel (growth factor reduced, Corning cat. #356231) were plated per well along the perimeter of each well of a 6-well plate. For passaging, organoids were incubated with Dispase (1 mg/ml, Gibco cat. #17105-041) for two hours at 37 °C. Pelleted organoids were treated with TrypLE Express (Gibco cat #12605-010) for five minutes at 37 °C with periodic pipetting to generate single cells for replating. Cultures were propagated for >10 generations before subcutaneous injection in mice.

Blood for circulating tumor DNA isolation was drawn into Streck tubes as previously described [39]. Plasma was isolated using a two-spin protocol: first for 20 minutes at 300 × g at 22 °C, and the upper plasma layer was transferred for a second round of centrifugation at 5,000 × g at 22 °C for 10 minutes. The buffy coat was retained separately.

### Patient-derived xenografts

Six-to seven-week-old male NOD *scid* gamma (NOD.Cg-*Prkdc^scid^ Il2rg^tm1Wjl^*/SzJ, NSG) mice were obtained from the Frederick National Laboratory for Cancer Research. Animal care was provided in accordance with the procedures and protocols approved by the NIH Animal Care and Use Committee. Pelleted organoid cells were rinsed and resuspended in HCM [14] containing 50% Matrigel, and 2 × 10^6^ cells in 100 µl were injected subcutaneously into the flank of NSG mice.

Bioluminescent PDX cells were generated as previously described [15]. Briefly, PDX tumors were isolated from mice and dissociated into single-cell suspension. Next, cells were transduced with firefly luciferase lentivirus and then cultured in Matrigel under puromycin (1 µg/mL) selection for one week, followed by a second round of growth and puromycin selection. Stable luciferase expression was confirmed by bioluminescent imaging (BLI) using the Xenogen BLI system.

To establish metastasis models, luciferase-expressing PDX tumors were removed from NSG mice and dissociated to single cell suspensions using gentleMACS Dissociator (Miltenyi) as previously described. 1 × 10^5^ cells were inoculated into the left cardiac ventricle of male NSG mice under 1.5% isoflurane anesthesia. The development and progression of metastases were monitored by weekly BLI. Mice were checked daily to observe signs of morbidity and euthanized following cachexia and/or paralgesia.

### Nucleic acid extraction and sequencing

From diagnostic biopsy tissues, genomic DNA and total RNA were extracted from formalin-fixed paraffin-embedded tissue sections mounted on slides, after macrodissection to enrich for viable tumor content using the AllPrep DNA/RNA FFPE Kit (Qiagen). For exome sequencing, after passing quality control steps, 100 ng of double-stranded genomic DNA extracted from tumor or benign FFPE slides was used for library preparation with the Nextera Flex for Enrichment assay with Exome Panel (Illumina). Libraries were pooled for target coverage of 50 × for benign DNA and 150 × for tumor DNA and sequenced on the NextSeq 550Dx instrument (Illumina). For RNA-seq, after passing quality control steps, 150 ng of total RNA extracted from tumor FFPE slides was used for library preparation with the TruSeq RNA Access Library RNA Exome Kit (Illumina). Libraries were sequenced on the NextSeq 550DX instrument (Illumina).

From blood, gDNA was extracted from 10–100 μL of buffy coat using the DNeasy Blood and Tissue Kit (Qiagen) following the manufacturer’s protocol with two elutions of buffer AE (100 μL per elution). Circulating tumor DNA (ctDNA) was extracted from plasma using the QIAamp Circulating Nucleic Acid Kit (Qiagen) following the manufacturer’s protocol with minor modifications: buffer ACB was incubated with lysate for 10 minutes on ice and elution was performed twice with buffer AVE (25 μL per elution) following a five-minute incubation at room temperature. ctDNA was processed into Illumina-compatible short-read sequencing libraries using the HyperPrep Kit (KAPA) without further fragmentation and sequenced on a 10B flowcell on a NovaSeq X Plus instrument (Illumina).

For NCI-LYM-1 tumor RNA sequencing, PDX tumor tissue was isolated from mice and dissociated into a single-cell suspension. Cells were processed using Mouse Cell Depletion Kit (Miltenyi) and human cells were recovered in PBS. RNA was extracted using the RNease Plus Mini Kit (Qiagen). Strand-specific, paired-end sequencing libraries were constructed using the Stranded mRNA Prep Kit (Illumina) and sequenced on a NovaSeq 6000 instrument (Illumina).

For NCI-LYM-1 tumor DNA sequencing, an aliquot of buffy coat from the donor was used as a reference sample. Both tumor cells and buffy coat were processed using the SP-GL2 kit for ultra-high molecular weight DNA isolation (Bionano). Aliquots of ultra high-molecular weight DNA were labeled using the DLS-G2 kit (Bionano), loaded onto G3.3 chips (Bionano) and imaged using the Saphyr instrument (Bionano). Additional aliquots of ultra-high molecular weight DNA were assembled into long-read sequencing libraries using the Ultra-Long DNA Sequencing Kit (cat. #SQK-LSK114, Oxford Nanopore Technologies, ONT) with a PCR-free protocol and sequenced on PromethION flowcells (cat. #FLO-PRO114M, ONT). Finally, additional aliquots of ultra-high molecular weight DNA were sheared with a Megaruptor shearing device (Diagenode) to 250–400 bp, processed into Illumina-compatible short-read sequencing libraries using the HyperPlus Kit (KAPA), and sequenced on a 10B flowcell on a NovaSeq X Plus instrument (Illumina).

### DNA analyses

Prior whole-exome sequencing (WES) data from LuCaP PDX models [40] was obtained from SRA via accession ID SRP008162, and prior whole-genome sequencing (WGS) data from mCRPC tumors [41, 42] was obtained from dbGaP via accession ID phs001648. Prior whole-exome sequencing from LuCaP PDX-O models and whole-genome sequencing from NCI PDO models were previously reported [14, 16].

From raw FASTQ files, sequencing adaptors were removed using trimmomatic (version 0.39) and reads were aligned to the human genome hg38 assembly using Burrows-Wheeler Aligner (BWA, version 0.7.17). Mapped reads were then coordinate sorted, and duplicates identified using MarkDuplicatesSpark (GATK, version 4.2.6.1), NM, MD, UQ tags fixed using SetNmMdAndUqTags (Picard), and recalibrated for base quality using BQSRPipelineSpark (GATK). Reads from multiple flow cells and lanes were merged and sorted for coordinates and marked for PCR duplicates again using MarkDuplicates. Averaged genome coverage and insert size were estimated using CollectWgsMetrics (WGS) or CollectHsMetrics (WES) and CollectInsertSizeMetrics (Picard).

For germline variant detection, Haplotypecaller (GATK, version 4.2.6.1) was used with ERC set as GVCF and a dbsnp138.vcf file used. Variant quality was assessed using VariantRecalibrator (GATK) for SNPs and indels independently with the following settings. For SNPs: --resource: hapmap, known=false, training=true, truth=true, prior=15.0 hapmap_3.3.hg38.vcf.gz --resource: omni, known=false, training=true, truth=false, prior=12.0 1000G_omni2.5.hg38.vcf.gz --resource: 1000G, known=false, training=true, truth=false, prior=10.0 1000G_phase1.snps.high_confidence.hg38.vcf.gz --resource: dbsnp, known=true, training=false, truth=false,prior=2.0 Homo_sapiens_assembly38.dbsnp138.vcf --an QD -an MQ -an MQRankSum -an ReadPosRankSum -an FS -an SOR -an DP --max-gaussians 4 -mode SNP. For indels: --resource: mills, known=false, training=true, truth=true, prior=12.0 Mills_and_1000G_gold_standard.indels.hg38.vcf.gz --resource: dbsnp, known=true, training=false, truth=false, prior=2.0 Homo_sapiens_assembly38.dbsnp138.vcf -an QD -an DP -an FS -an SOR -an ReadPosRankSum -an MQRankSum --max-gaussians 4 -mode INDEL. The detected germline variants were then recalibrated using ApplyVQSR (GATK) with the quality scores determined and ts-filter-level set to 99.5 for SNPs and 99.0 for indels. To identify the functional impact of the detected variants, ANNOVAR (version 2020-06-08) was used for variant annotation. First, the genotype calling format was converted to ANNOVAR format using convert2annovar.pl with the following settings: -format vcf4 --keepindelref -include -withzyg. Second, variants were annotated using table_annovar.pl with the following settings: --tempdir temp --thread 4 --buildver hg38 --remove --otherinfo --protocol refGene, cosmic70, gnomad_exome, avsnp150 --operation g,f,f,f --nastring”.

For ancestry inference, we extracted genotypes from the germline whole-genome BAM at 29 African ancestry-informative markers described previously [43] using bcftools, retaining sites with depth ≥6 and genotype quality ≥10. We merged the patient with 1000 Genomes Phase 3 reference individuals (AFR, EUR, AMR) at these loci and formatted diploid genotypes for STRUCTURE. Unsupervised analyses (K = 3) were performed for quality control; final estimates used supervised STRUCTURE (USEPOPINFO = 1) with AFR and EUR references designated as training populations (K = 2), using 100,000 burn-in and 100,000 MCMC iterations, repeated in 10 runs. We reported the mean cluster membership (Q) across runs for the patient.

For somatic variant detection, Mutect2 (GATK, version 4.2.6.1) was used. A panel of normal (PON) was generated by calling variants in buffy coat samples using Mutect2 in a tumor-only mode (parameters set to: --disable-read-filter MateOnSameContigOrNoMappedMateReadFilter -max-mnp-distance 0 --native-pair-hmm-threads 24), GenomicDBImport (--merge-input-intervals true), and CreateSomaticPanelOfNormals. Read contamination was calculated using GetPileupSummaries and CalculateContamination. Somatic variants were then detected using Mutect2 with the following settings: -pon the PON generated --germline-resource af-only-gnomad.hg38.vcf.gz --af-of-alleles-not-in-resource 0.0000025 --native-pair-hmm-threads 24 -A StrandBiasBySample --disable-read-filter MateOnSameContigOrNoMappedMateReadFilter. Read orientation bias was applied using LearnReadOrientationModel and variants were filtered using FilterMutectCalls with the read contamination and orientation bias previously calculated. Finally, somatic variants were annotated using ANNOVAR with the same settings as described for germline variants.

For somatic copy number variant (CNV) detection, a composite of GATK toolsets were used. Read depths and allelic counts were measured using CollectReadCounts (--interval-merge-rule OVERLAPPING ONLY) and CollectAllelicCounts, respectively. The resulting hdf5 file from the buffy coat samples were used to create a panel of normals using CreateReadCountPanelOfNormals. Background signals were removed from tumor read depths using DenoiseReadCounts. Normalized copy ratios were determined using ModelSegments which used the de-noised read depths and allelic ratios calculated previously (--number-of-changepoints-penalty-factor 5) and CallCopyRatioSegments. Genome-wide CNVs were visualized using Integrated Genome Viewer. In addition, titanCNA [44] was used to model copy number variation when matched control was available.

For somatic structural variant (SV) detection, a copy number-sensitive SV-calling algorithm set, GRIDSS/LINX [45], was used. Read depths and SNP allelic frequency were measured using COBALT (version 1.13; -gc_profile GC_profile.hg38.1000bp.cnp) and AMBER (version 3.9; -ref_genome_version V38 -loci 1000G_phase1.snps.high_confidence.hg38.vcf.gz), respectively. SVs were called using GRIDSS (version 2.13.2; --steps preprocess,assemble,call) and somatic SVs filtered using GRIPSS (version 2.3.2; -ref_genome-version 38). PURPLE (version 3.1; -amber AMBER output -cobalt COBALT output -gc_profile GC_profile.hg38.1000bp.cnp -ref_genome_version 38 -structural_vcf gripss.filtered.vcf.gz -sv_recovery_vcf gripss.vcf.gz -somatic_vcf Mutect2 vcf) was used to estimate tumor ploidy and copy number profile. SV clusters and potential driver events including gene fusions were then determined using LINX (version: 1.22; -ref_genome_version 38 -sv_vcf purple.sv.vcf.gz - purple_dir PURPLE output -check_fusions –known_fusion_file known_fusion_data.38.csv - ensembl_data_dir ensembl_data -check_drivers -driver_gene_panel DriverGenePanel.38.tsv - fragile_site_file fragile_sites_hmf.38.csv -line_element line_elements.38.csv).

For transcriptional activity inference from circulating tumor DNA, Griffin was used [46]. GC ratio and sequence mappability were determined using griffin_GC_and_mappability_correction.snakefile and nucleosome occupancy was then quantified at each transcription factor binding sites using griffin_nucleosome_profiling.snakefile.

For phylogenetic reconstruction, both PhylogicNDT [47] and a cfDNA-sensitive toolset [18] were used. The clonal prevalence of each functionally annotated somatic variant identified by Mutect2 was determined using ABSOLUTE. Tumor clonal structures were then inferred using the PhylogicNDT toolset. For the second clonal inference algorithm, variants that were present in both PDX and cfDNA were jointly identified following the same Mutect2 pipeline described above. Variants supported by at least eight counts and whose allelic frequency was greater than ten times of that from the buffy coat were collected. Heterozygous SNPs and log copy ratios were also used to model clonality. Tumor ploidy and truncal copy number model were determined using copy number segments, and the subclones truncal model and tumor clonality were inferred using subclonal clusters.

Base calling in long-read sequencing POD5 files was performed using MinKNOW version 23.04.6 and Guppy version 6.5.7. An estimate of a hundred-fold genome coverage was supported. Sequence read length N50 of at least 20 kb suggested high genome coverage with predominantly long reads. Sequence reads were then mapped to the human genome (GRCh38) using minimap2 (version 2.26; -ax map-ont). Clair3 (version: 1.0.4; --platform=ont --model_path= r1041_e82_400bps_hac_v420 --enable_phasing --longphase_for_phasing) was used to identify germline variants from the buffy coat which were used to phase long reads using whatshap (version: 1.1; --ignore-read-groups --tag-supplementary). Phased reads were then used to identify somatic small variants using ClairS (version: 0.1.6; run_clairs --platform ont_r10_guppy) and structural variants using severus.py (version: 0.1.1; -vntr-bed human_GRCh38_no_alt_analysis_set.trf.bed), sniffles (version: 2.2), and nanomonsv. For 5-methylcytosine (5mC) and 5-hydromethylcytosine (5hmC) base calling, POD5 files were separately processed using dorado base caller (version: 0.8.2; --device cuda:all –min-qscore 9 dna_r10.4.1_e8.2_400bps_hac@v4.2.0 --modified-bases 5mCG_5hmCG). The resulting reads were then mapped to the human genome hg38 assembly using dorado aligner (version: 0.8.2) and coordinate sorted using samtools (version: 1.21). DNA methylation per CpG site was quantified using modkit pileup (version: 0.4.1; --cpg) and methylkit was used to perform differential methylation analysis. Experimentally validated transcription factor binding sites (TFBSs) were obtained using GTRD [48] and used to identify TFBSs that were differentially methylated.

For optical genome mapping, signals from Saphyr images were converted to a collection of molecules mapped to the human genome followed by de novo assembly of genome maps. Rare variant analysis was then performed and structural variants supported by a variant allele frequency greater than 0.1 were detected and viewed using Bionano Access.

### RNA analyses

Prior RNA sequencing from mCRPC [42] was obtained from dbGaP via accession ID phs001648. Prior whole-transcriptome sequencing data from LuCaP PDX and NCI PDO models were previously reported [16, 49]. For PDX tumors, reads that preferentially mapped to the murine genome (mm10) were removed using Disambiguate and the rest of sequence reads, expected to be primarily human reads, were mapped to the human genome assembly hg38 using tophat (version 2.1.2; --min-anchor-length=10 --num-threads=12 --b2-sensitive --no-novel-juncs). Gene counts were calculated using rsem-calculate-expression (STAR 2.7.11b, RSEM 1.3.3; --star --paired-end --estimate-rspd -p 30 --calc-ci --ci-memory 8192). For principal component analysis (PCA), raw gene counts normalized by variance stabilizing transformation (VST) were used, principal components (PCs) defined and weighted, and transcriptomic similarities were determined between samples. To evaluate pathway enrichment across tumors and PDX/PDO models, VST-normalized gene counts were used for gene set variation analysis [50] using the mSigDB hallmark gene set. Alternatively, normalized gene counts were used to perform differential gene expression analysis using DESeq2. Statistically significant genes with a P value less than 0.05 were used for pathway analysis. Pathway enrichment using the upstream regulator module was performed using Ingenuity Pathway Analysis (releases 2024 Q3 through 2025 Q3). Candidate upstream regulators were filtered for genes, pathways, and transcription factors and sorted by bias-adjusted z-score, with or without false discovery correction using the Benjamini-Hochberg method. Up to 200 genes (150 upregulated, 50 downregulated) were used to identify potential drug sensitivities using the Connectivity Map [51].

### Histopathology

Bony tissues (long bones and spine) and soft tissues (adrenal, kidney and liver) were harvested when mice reached ethical end points. Tissues were fixed in 4% paraformaldehyde, replaced with 70% ethanol solution after 24 hours, and paraffin embedded for sectioning. Long bones and spine were decalcified in 10% ethylenediaminetetraacetic acid before embedding. Bone sections were stained with hematoxylin and eosin (H&E) and orange G, and soft tissues were stained with H&E. Bone sections were stained with tartrate-resistant acid phosphatase (TRAP) for determining the number of osteoclasts [52]. Masson’s Trichrome stain (Abcam, cat. #ab150686) was used to visualize new bone formation through collagen-containing osteoid in bone.

For immunodetection (colorimetric or fluorescent), paraffin sections were cut at five micron thickness onto charged slides. Tissue sections were deparaffinized in xylene and rehydrated through graded ethanol washes. Heat induced epitope retrieval (HIER) was processed using a Decloaking Chamber (Biocare Medical) for 15 minutes at 110 °C and 5 psi. For anti-AR (Cell Signaling, cat. #5153), anti-SOX2 (Cell Signaling, cat. #3579), anti-ASCL1 (Cell Signaling, cat. #10585), anti-Ki-67 (Cell Signaling, cat. #9027), anti-E-cadherin (Cell Signaling, cat. #3195) and anti-cytokeratin 8 (Novus Biologicals, cat. #NBP2-16094), HIER was performed in Diva Decloaker pH 6 (Biocare Medical, cat. #DV2004MX). For anti-SYP (Dako, cat. #M7315) HIER was performed in Tris-EDTA pH 9 (Abcam, cat. #ab93684). After HIER, slides were loaded into an intelliPATH FLX automated slide stainer (Biocare Medical). Slides were blocked for endogenous peroxidases for 10 minutes with 3% H_2_O_2_ (Fisher Scientific, cat. #M-15590) and blocked for background protein for 30 minutes with Background Punisher (Biocare Medical, cat #BP974MM). Primary antibody was diluted at 1:200 (AR, SOX2, ASCL1, SYP), 1:250 (cytokeratin 8), 1:400 (E-cadherin), or 1:500 (Ki-67) Renoir Red diluent (Biocare Medical, cat. #BMPD904M) and incubated with tissues for 60 minutes. For IHC, tissues were subsequently incubated for 30 minutes with MACH 4 Universal HRP-Polymer (Biocare Medical, cat. #M4U534) for rabbit primary antibodies, adding MACH 4 Probe reagent for mouse primary antibodies. Colorimetric detection was achieved using Betazoid DAB (Biocare Medical, cat. #BDB2004) for five minutes, counterstained with CAT Hematoxylin (Biocare Medical, cat. #CATHE-M), dehydrated through graded alcohols, cleared in xylenes, and mounted with Permount Mounting Medium (Fisher Scientific, cat. #SP15). For IF, tissues were subsequently incubated for 30 minutes in 1:250 dilutions of Alexa Fluor-conjugated secondary antibodies (Invitrogen, cat. #A21245, cat. #A11004, and cat. #11006) and mounted using DAPI-antifade solution. All slides were scanned at high resolution on Carl Zeiss Axioscan.Z1 microscope using a ×20 Plan-Apochromat (NA 0.8) objective (Zeiss) tuned for brightfield or fluorescent settings.

Nuclear (Ki-67, AR, SOX2, and ASCL1) and cytoplasmic (SYP) staining was determined for PDX tissues by a pathologist (S.K.). For AR, SOX2, ASCL1 and SYP, tumor staining was measured using the H-score method, a semiquantitative assessment of staining intensity that reflects antigen concentration. H-score was determined according to the formula: ([% of weak staining] × 1) + ([% of moderate staining] × 2) + ([% of strong staining] × 3), yielding a range from 0 to 300. For Ki-67, the fraction of tumor cells with any positive nuclear signal was calculated to determine the Ki-67 index, yielding a range from 0 to 100.

### In vitro drug sensitivity

Two thousand NCI-LYM-1 cells were seeded per well of a flat, white bottom 384-well plate in 20 µl of 70% Matrigel to form organoids. 30 µl HCM was added one hour after solidification. 24 hours later, up to 10 µl of test compounds (dissolved in DMSO) or DMSO were dispensed into each well based on a 10-point concentration range (0.05–30 µM) using a Tecan D300e Digital Dispenser with two-fold serial dilutions. Organoid drug testing was performed with carboplatin (Selleckchem, cat. # S1215), docetaxel (Selleckchem, cat. #S1148), topotecan, navitoclax, AZD-5991, berzosertib, talazoparib, and ipatasertib (all from the NCI Developmental Therapeutics Program Open Chemicals Repository). Organoids were retreated with fresh media containing drug every two days. After seven days, cell viability was measured using the CellTiter-Glo 3D Assay (Promega) following the manufacturer’s protocol, with luminescence measured using a GloMax microplate reader (Promega). Luminescence activity was normalized to the average intensity of the DMSO control. Dose-response curves were generated by fitting the normalized viability data to a three-parameter logistic regression model with variable slope constrained from 0–100% viability using GraphPad Prism version 9. All assays were performed independently three times with three technical replicates for each dilution.

### Statistics

Statistical analyses were performed with GraphPad Prism version 10, Microsoft Excel for Mac version 16, and R version 4.4.2 (R Core Team 2023). Comparison of single factors between de novo and transformed tumors was performed using Welch’s *t* tests. Comparisons of covariates within single factors between de novo and transformed tumors were performed using Fisher’s exact test. The application of clinical or pathologic variables to clinical outcomes was measured using Kaplan-Meier survival estimates with the log-rank test for radiographic progression. Pathway enrichment and drug perturbation was filtered and sorted by *P* values, adjusted *P* values and bias-corrected *z* scores.

## Data Availability

All data produced in the present work are contained in the manuscript.

## DECLARATIONS

### Ethics approval and consent to participate

The collection and analysis of tissue from patients with metastatic prostate cancer was approved by the Institutional Review Board of the National Institutes of Health (NCT04826341, “A phase I/II study of sacituzumab govitecan plus berzosertib in small cell lung cancer, extra-pulmonary small cell neuroendocrine cancer and homologous recombination-deficient cancers resistant to PARP inhibitors”, PI: Anish Thomas). All patients provided written informed consent before participating. This research was conducted in accordance with the principles of the Declaration of Helsinki.

### Consent for publication

Not applicable.

### Data availability

The data underlying this article have been deposited in Database of Genotypes and Phenotypes (dbGaP) and Gene Expression Omnibus (GEO) at https://www.ncbi.nlm.nih.gov/gap/ and https://www.ncbi.nlm.nih.gov/geo/, respectively, and can be accessed with phs001587.v4.p1 (dbGaP), phs001813.v4.p1 (dbGaP).

### Materials availability

The NCI-LYM-1 model is available to researchers upon request.

### Authors’ contributions

<u>Data acquisition</u>: C.L., J.Y., K.B., J.T., T.T., D.V.

<u>Methodology</u>: C.L., J.Y., S.K., K.K.

<u>Reagents</u>: J.Y., C.C.

<u>Patients</u>: M.A., S.N., K.P., A.T.

<u>Analysis</u>: C.L., K.B., S.K., C.C.

<u>Manuscript:</u> All authors

<u>Supervision</u>: A.G.S., W.D.F., A.T.

<u>Accessed and verified all underlying data</u>: A.G.S.

## Acknowledgments

The authors gratefully acknowledge the patients and the families of patients who contributed to this study.

This research was supported in part by the Intramural Research Program of the National Institutes of Health (NIH) and is subject to the NIH Public Access Policy. Through acceptance of this federal funding, the NIH has been given a right to make the work publicly available in PubMed Central. The contributions of the NIH authors are considered Works of the United States Government. The findings and conclusions presented in this paper are those of the authors and do not necessarily reflect the views of the NIH or the U.S. Department of Health and Human Services.

Portions of this work utilized the computational resources of the NIH HPC Biowulf cluster.

