## Supplementary Figure 1 for "Molecular decoupling of lineage identity and morphology in aggressive variant prostate cancer"

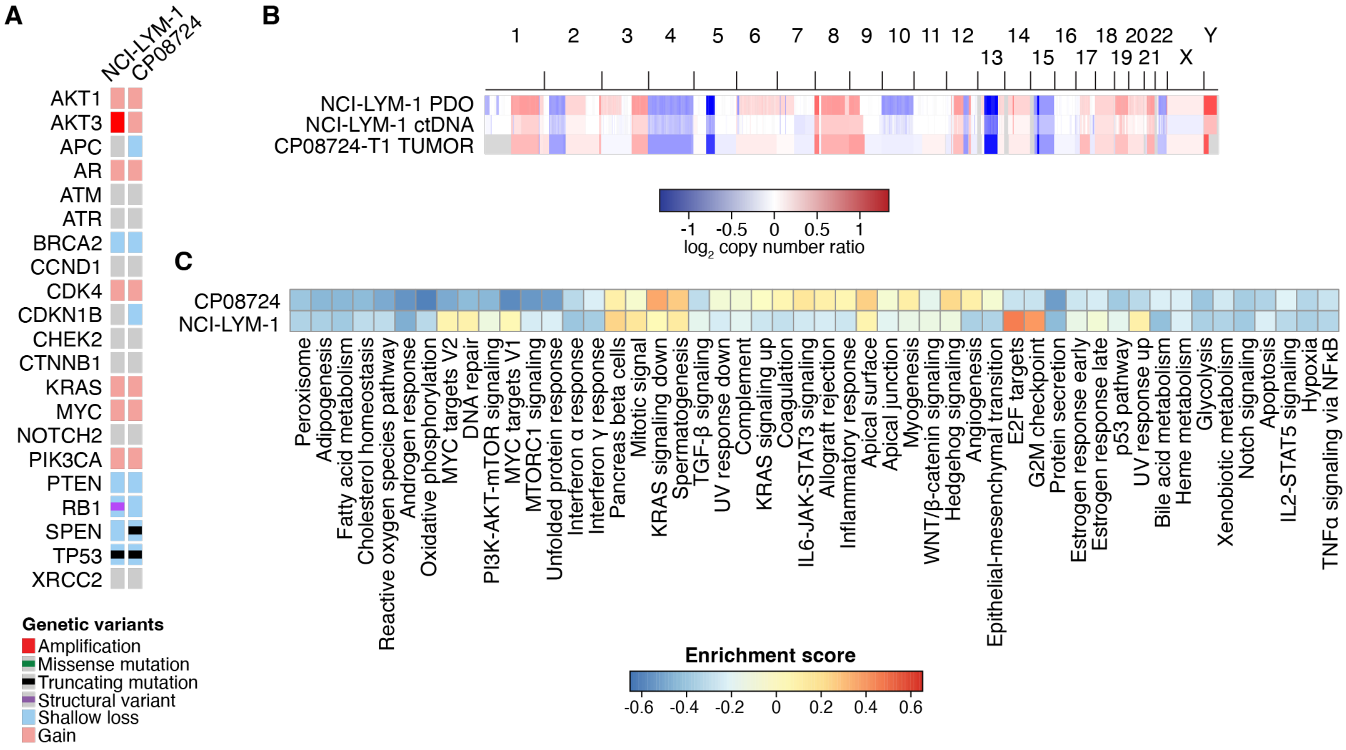


**Supplementary Figure 1. Comparison of molecular features between NCI-LYM-1 and donor tumor biopsy. (A)** Oncoprint depicting copy number and mutational status of selected prostate cancer genes NCI-LYM-1 and donor tumor biopsy. All mutations shown were curated for known oncogenic status. **(B)** Whole-genome somatic copy-number estimates derived from whole-genome sequencing NCI-LYM-1 and whole-exome sequencing of donor tumor biopsy. **C)** Heatmap depicting unsupervised nonparametric gene set variation analysis for NCI-LYM-1 and donor tumor biopsy transcriptomes projected against the mSigDB Hallmarks gene sets.
